# No excess mortality detected in rural Bangladesh in 2020 from repeated surveys of a population of 81,000

**DOI:** 10.1101/2021.05.07.21256865

**Authors:** Prabhat Barnwal, Yuling Yao, Yiqian Wang, Nishat Akter Juy, Shabib Raihan, Mohammad Ashraful Haque, Alexander van Geen

**Affiliations:** Department of Economics, Michigan State University, USA; Department of Statistics, Columbia University, USA; Innovations for Poverty Action, Bangladesh; Lamont-Doherty Earth Observatory, Columbia University, USA

## Abstract

**Background:** Excess mortality has demonstrated under-counting of COVID-19 deaths in many countries but cannot be measured in low-income countries where civil registration is incomplete.

**Methods:** Enumerators conducted an in-person census of all 16,054 households in a sample of 135 villages within a 350 km^2^ region of Bangladesh followed by a census conducted again in May and November 2020 over the phone. The date and cause of any changes in household composition, as well as changes in income and food availability, were recorded. For analysis, we stratify the mortality data by month, age, gender, and household education. Mortality rates were modeled by Bayesian multilevel regression and the strata aggregated to the population by poststratification.

**Results:** A total of 276 deaths were reported between February and the end of October 2020 for the subset of the population that could be contacted twice over the phone, slightly below the 289 deaths reported for the same population over the same period in 2019. After adjustment for survey non-response and poststratification, 2020 mortality changed by -8% (95% CI, -21% to 7%) relative to an annualized mortality of 6.1 per thousand in 2019. However, salaried breadwinners reported a 40% decline in income and businesses a 60% decline in profits in May 2020.

**Discussion:** All-cause mortality in the surveyed portion of rural Bangladesh was if anything lower in 2020 compared to 2019. Our findings suggest various restrictions imposed by the government limited the scale of the pandemic, although they need to be accompanied by expanded welfare programs.

**Key questions:** *What is already known?:* Civil registry data from dozens of countries, where available, indicate gaps between official death counts attributed to COVID-19 and, usually, a larger increase in total mortality in 2020 compared to previous years. This approach is not available to gauge the impact of COVID-19 in countries such as Bangladesh where the civil registry system is slow and coverage incomplete. One year after the first COVID-19 case was reported in Bangladesh in 2020, the number of deaths attributed to COVID-19 was equivalent to 1% of annual mortality in previous years. Whether this low figure compared to many other countries is an accurate reflection of the situation or is distorted by massive under-counting has been much debated, albeit on the basis of little direct evidence. The lack of accurate mortality data has made it only more difficult for policy makers to balance the public health benefit of lockdowns and similar measures relative to the well-documented economic costs and hardship imposed by such measures on poor households in particular. A PubMed search conducted on May 4, 2021 under (Bangladesh[Title/Abstract]) AND (excess mortality[Title/Abstract]) limited to 2020-21 did not yield a single relevant study.

*What are the new findings?:* By conducting of repeated census of a large rural population over the course of 2020, once in person and twice over the phone, we document if anything a slight decline in mortality across a rural area of Bangladesh compared to 2019. We also place an upper limit on the level of under-reporting at the national level that is consistent with our observations. At the same time, interviewed households reported a large and sustained drop in income as well as reduced access to food.

*What do the new findings imply?:* The impact of the pandemic on mortality was thankfully limited in rural study area of Bangladesh in 2020. This suggests that officially recorded COVID-19 deaths may have been contributed largely by the urban population, about a third of the country overall. At the same time, the economic and nutritional impact of restrictions on trade and movement was substantial and probably underestimated in the rural population. As cases surge again, as they did in March–April 2021, policy makers may want to consider limiting strict restrictions to urban areas while expanding a financial support throughout the country.

## INTRODUCTION

Excess mortality has received increasing attention over the course of the pandemic as a robust measure of the net impact of COVID-19, especially when comparing countries with different reporting systems and different definitions of COVID-19 deaths (1). The approach has revealed under-reporting of COVID-19 deaths by an order of magnitude in several countries (2). The key assumption underlying this approach is that mortality data are reliably compiled within a reasonable time frame through a country’s civil registration system, however. This is not the case in most low-income countries where, therefore, excess mortality cannot be reliably measured and under-reporting of COVID-19 deaths is particularly difficult to assess (3–5).

The lack of reliable excess mortality data in low-income countries is a serious issue for at least two reasons. First, citizens, policy makers, and donor agencies cannot know the actual impact of COVID-19 without reliable mortality data and act accordingly (6). Second, recent surveys conducted across several low-income countries have shown that the poor have been particularly hurt economically by the pandemic (7). Lack of reliable mortality data makes it even more difficult for policy makers to gauge the trade-off between the economic costs and public health benefits of a blanket lockdown or other more targeted restrictions.

During the year that ended March 1, 2021, a total of 8,400 deaths in Bangladesh had been officially attributed to COVID-19 (8). This corresponds to 1% of the 820,000 annual deaths calculated for a total population of 167 million and an annual mortality rate of 4.9 per 1000 estimated from the government’s sample vital statistics data (9). However, only a third of deaths are currently officially recorded within 45 days in Bangladesh (10). Considering the high excess mortality estimated from burial records in some countries without a functioning civil registry, such as Indonesia (11) and Yemen (12) for instance, the actual number of COVID-19 deaths could be considerably higher than the national data suggest. The possibility of under-reporting of a first wave of COVID-19 cases and fatalities in South Asia has been widely discussed but on the basis of limited direct evidence (5,13,14). The goal of this study was determine whether the low number of official COVID-19 deaths in Bangladesh in 2020 could have been the result of massive under-counting. The issue is salient for policy makers in Bangladesh as the COVID-19 cases and mortality increased rapidly starting in early March 2021.

According to government data, a majority of COVID-19 cases confirmed by rRT-PCR testing have been concentrated in the capital Dhaka and the next largest city Chattogram (figure 1). Thousands of cases have also been reported from more remote districts, however. The pattern leaves no doubt that the pandemic has spread across the entire the country. The low number of official COVID-19 deaths therefore cannot be attributed to effective isolation of the most affected areas from the rest of the country. The first confirmed cases of COVID-19 were reported on March 8, 2020. In spite of a de facto lockdown declared on March 23, the number of confirmed cases rose rapidly to reach a maximum over 20,000 per week in June 2020 (figure 2a). Over much of the summer, the number of deaths attributed to COVID-19 hovered around 250 per week. Both cases and deaths then gradually declined through February 2021, with the exception of a temporary resurgence in November 2020. Since early March 2021, both COVID-19 cases and mortality have increased rapidly to levels above their peaks reached in June 2020.

**Figure 1.**
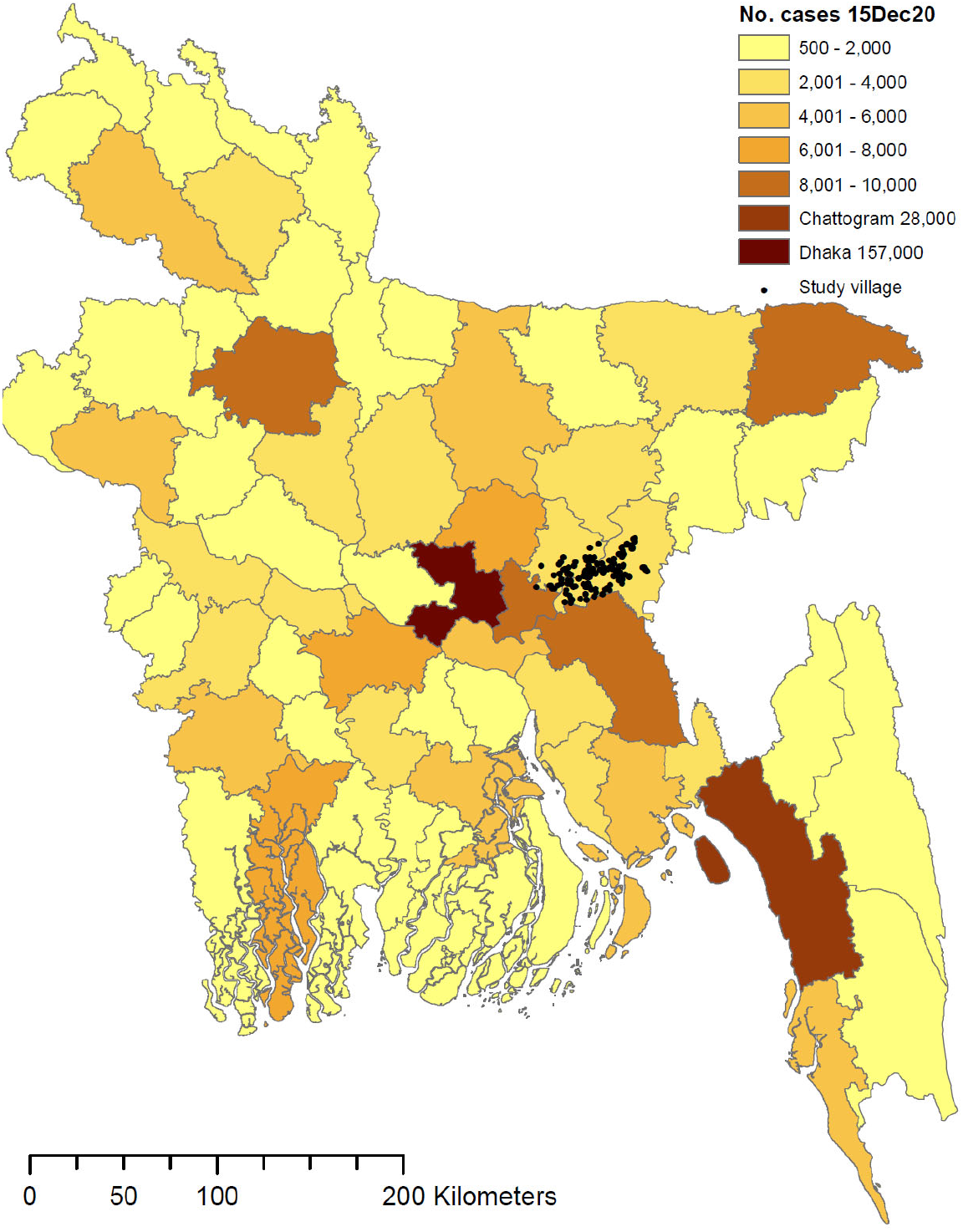
Map of Bangladesh showing the distribution of confirmed COVID-19cases as of December 15, 2020. Study villages surveyed in person and over the phone in 2020 are shown as black dots east of the capital Dhaka.

**Figure 2.**
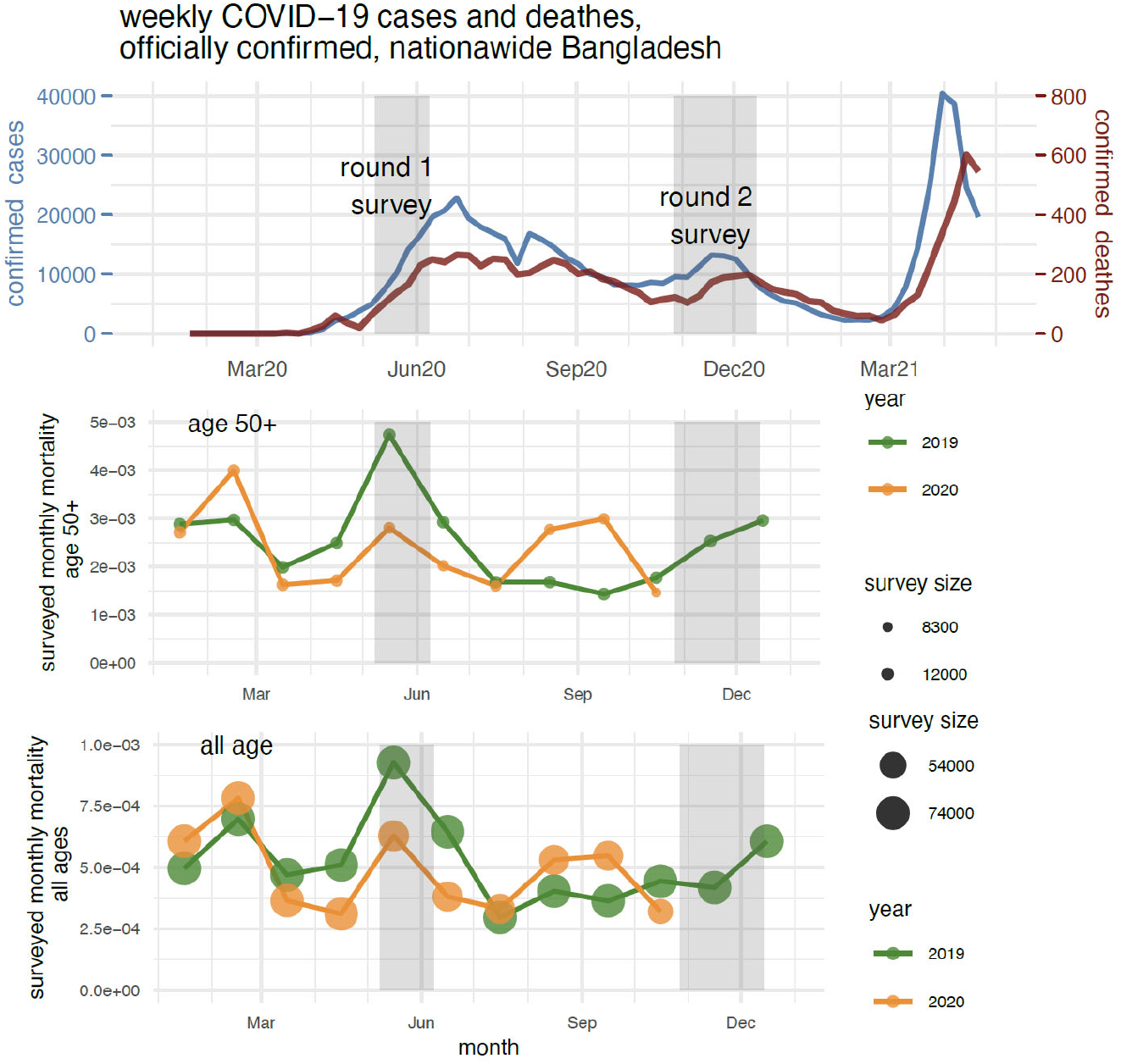
(a) Weekly time series of confirmed COVID-19 cases (blue curve) and deaths (red curve) in Bangladesh in relation to timing of our phone surveys, from February 2020-April 28, 2021. (b, c) Monthly sample mortality rate for elderly (age 50+) and all ages in 2019 and 2020 in study villages. The mortality rate is calculated relative to the size of the survey sample for that month, which is indicated by the dot size.

Using detailed survey data containing individual level information for a sizeable population, we estimate here month-, age-, gender-, and education-specific mortality. We calculate excess mortality relative to 2019 after adjusting for key characteristics and averaging over the January 2019 census population. We complement this analysis with data on the economic impact of the pandemic obtained during the same phone surveys.

## METHODS

### Census survey

Using an approach developed for conflict zones (15), we inferred mortality in our study area in 2020 by repeatedly conducting a census of a large number of households, once in person in early 2020 and two more times over the phone later in the year. The in-person census involved a team of 60 enumerators working in the field for two weeks; each of the phone surveys employed full-time another 60 enumerators for an entire month. At the beginning of each phone call, each consenting respondent was asked to list current household members without prompting. The electronic questionnaire was set up for the enumerator to check off each household member recorded in January 2020 and identify discrepancies to investigate subsequently. As a reference for identifying excess mortality in 2020, each household was asked at the end of the first phone census about all deaths that occurred in 2019. Only for 2019 was a respondent asked directly about a death that occurred in the household.

Repeated census surveys were conducted in 135 villages, or pre-defined portions (“paras”) of larger villages, located 30–100 km to the northeast of Dhaka (figure 1). The villages had been selected for a randomized controlled trial, paused in February 2020, to reduce arsenic exposure from drinking well-water. The 135 study villages are distributed across 16 unions, an administrative unit of which there are a total of 4,563 in Bangladesh. Government data show that the average age in these 16 unions is 3 years higher than in the 2,188 other rural unions of the country and that the proportion of households engaged in agricultural activity is somewhat lower (Table 1). Proxies for socioeconomic status such as education and the number of rooms in the house are no different in the study unions compared to other rural unions.

**Table 1.** Socioeconomic statistics in rural Bangladesh nationwide from the 2016 household income and expenditure survey(16) (distributed across 2,204 unions), and in our sample survey. The number of observations (n) corresponds to individuals for age (years) and sex (1=Female, 0=Male), and to households for education (1=over half of household members received any formal education), main economic activity (proportion of household members engaged in agricultural sector), and count of rooms in the house.

| Variable |  | Unions in rural Bangladesh |  |  | Unions in this study |  |
| --- | --- | --- | --- | --- | --- | --- |
|  | N | Mean | SD | n | Mean | SD |
| Age | 129,347 | 28 | 19 | 1,394 | 25 | 19 |
| Sex | 129,348 | 0.501 | 0.500 | 1,394 | 0.507 | 0.500 |
| Education | 31,795 | 0.85 | 0.36 | 300 | 0.85 | 0.35 |
| Rooms in house | 31,758 | 2.3 | 1.3 | 300 | 2.4 | 2.5 |
| Agricultural activity | 31,795 | 0.43 | 0.47 | 300 | 0.26 | 0.41 |

From January 15 to February 3, 2020, enumerators sought to contact door-to-door all 17,538 households identified in the study villages or paras. Among these households, 1,478 were absent and could not be reached. Of the remaining 16,060 households, only 6 declined to participate in what was presented at the time as a study of arsenic mitigation. Sharing a kitchen was the criterion used to define a household. Following consent, the name, age, gender, and relationship of each individual member of the household, as well as GPS coordinates of each house, were recorded electronically. Up to two mobile phone numbers from all consenting households were also recorded. Most households were subsequently recontacted using one of these numbers, or in some cases alternative numbers provided by their neighbors. During the first phone survey conducted from May 8 to June 7, 2020, a total of 14,551 (91%) of the households surveyed in person in January 2020 could be contacted and consented to respond. During the second phone survey conducted between October 27 and December 14, 2020, 11,933 (74%) households consented. The total population of consenting households surveyed in person amounts to 81,164. This number is used as the reference population and includes 7,921 household members recorded during the phone surveys who had reportedly been overlooked during the January 2020 in person census, as well as 484 deaths in the households that occurred reportedly in 2019. The reference population does not include 1,068 household members who joined the household from elsewhere in 2020.

Additional information related to 2019 and 2020 deaths was collected during the phone surveys. Respondents were asked if a reported death was the result of injury, e.g. a road accident, and if treatment from a doctor or at a hospital was sought. Respondents were asked if death was preceded by symptoms related to COVID-19 such as fever, headache, cough, sore throat, breathing difficulty, loss of sense of smell, muscle aches, and chills. Respondents were also asked to attribute reported deaths to a few broad categories including stroke or heart disease (combined here because they are often confused in rural Bangladesh), cancer, liver, or lung disease. Some deaths, often in the case of an elderly parent, were reported more than once by different households. A total of 76 such cases were identified over 2019–2020 based on name and proximity and then confirmed with an additional phone call to avoid duplication. In order to determine the economic impact of COVID-19 in Bangladesh, we requested additional information from a randomly selected 20% of 16,054 households in the first phone survey and the same 20% with an additional 8% of households in the second phone survey. A total of 2,608 households participated in the first survey and 3,151 households in the second.

### Multilevel regression model of excess mortality and poststratification

We estimate mortality using a multilevel regression model and poststratification for two main reasons: first, to adjust for attrition i.e., non-response in census round 1 or 2, or both, and second, to understand the variation across demographic groups. The underlying assumption is that attrition is a function of age, gender and education. To estimate aggregate mortality, we therefore first estimate mortality for various age, gender, and education group and then calculate a weighted sum.

We stratify observed death counts into cells based on a four-way structure: (a) month (22 levels: Jan 2019, Feb 2019, …, Oct 2020), (b) age during the evaluation month (binned into 9 levels: 0–9 years old, 10–19, …, 70–79, 80 and above), (c) gender (2 levels, 1 = female), and (d) household education level (2 levels indicating if less (=1) or at least half (=2) of the adult members in the household have ever received some formal education.

In order to compare mortality before and during the pandemic, and to estimate the resulting excess mortality, we need to specify the local onset of impact of COVID-19 in our study villages. We primarily consider February 2020 as the start month but also consider later months. We specify a comparison-starting month *t*_start_, which splits time into the two periods (a) the baseline: Jan 2019, Feb 2019, …, *t*_start_ − 1, and (b) the potential excess period: *t*_start_, *t*_start_ + 1, …, Oct 2020. This approach allows us to be agnostic about when COVID-19 started to impact mortality in the study villages.

#### Modeling baseline and excess mortality in each strata

Both baseline and excess mortality may differ across strata. We set up a multilevel logistic regression (17,18) to model baseline mortality in each stratum by a four-way interaction of month-of-year, age, sex and education (Appendix A). To quantify the stratum-specific excess mortality (the additional risk in 2020 after month *t*_start_), we set another layer of multilevel model, where the monthly excess mortality is decomposed by a three-way interaction of age, sex and education.

#### Partial-pooling using Bayesian inference

We complete the model with a lag-1 autoregressive (AR) prior on all age effects and month effects, and weakly-informative priors on other coefficients. We perform fully Bayesian inference for our complete model in Stan using 4 chains and 3000 posterior simulation draws for each chain. (19) Computational diagnostics indicate all chains mix well.

#### Aggregating mortality and excess mortality

To aggregate the stratum-specific mortality to the population of interest, we generate posterior simulation draws of mortality in each stratum. Under the assumption that stratum-specific mortality rates are not affected by attrition in later rounds of census survey, the aggregate mortality is a weighted sum of mortality in each stratum, where weights are constructed using the January 2020 census data, from which we compute the excess mortality in the population by the Monte Carlo method. The poststratified excess mortality in equation [eq_excess_m] implicitly compares the averaged mortality in *t*_start_, *t*_start_ + 1, …, Oct 2020, with the same period in 2019. We repeat the same process to calculate the population-aggregated age-specific excess mortality change (Appendix A).

## RESULTS

### Raw data

Considering first only the 11,256 households that could each be reached during both phone surveys, a total of 639 deaths were reported between January 2019 and the end of October 2020 for a total population of 58,806, excluding individuals who joined the household after the January 2020 census. This corresponds to an average annualized mortality rate of 5.9 per 1000 over 22 months. For the same households that could be reached twice over the phone, a total of 276 deaths were reported between February and the end of October 2020, slightly below the 289 deaths reported for over the same months in 2019. Using all the available data, the monthly-sample-weighted annualized mortality rate between January 2019 and Oct 2020 is 6.1 per thousand (figure 2). The challenge is to determine the extent to which mortality differed before and after the pandemic by taking into account all the available data, including households that could not be reached after the January 2020 in-person census. Broken down by month, the annualized sample mortality rate in the study population varied between 3.5 and 11 deaths per 1000 since January 2019, but without a clear seasonal pattern (figure 2b). A longer time series from official statistics suggests that mortality in rural Bangladesh is on average 20% higher during winter compared to summer (20). The highest mortality rate during our study period was recorded in May 2019, but this could also be an artifact of the first phone survey being conducted in May 2020 and households reporting that a member died about a year ago. The data do not show an increase in mortality that coincides with the peak in COVID-19 cases and deaths reported centered on late May 2020 (figure 2a). The second round of phone surveys only partly covers the second peak in COVID-19 cases and deaths centered on November 2020. The age distribution of the surveyed population (figure 2) is comparable to published national trends (9). The number of deaths as a function age over the entire study period shows an initial drop beyond ages 0–5 followed by a steady increase in the number of deaths to age 80 and above, in spite a diminishing contribution to the overall population. Beyond these simple compilations, modeling is required to infer representative mortality trends.

The model confirms the expected increase in mortality as a function of age, expressed as a log odds ratio (figure 3a). There is no clear seasonal pattern in mortality in our data according to the model (figure 3b). Excess mortality is the key output from the model. At first, we assume that the onset of the pandemic in the study villages was in February 2020. The model shows no excess mortality during February–October 2020 and, if anything, possibly a slightly lower mortality log odds ratio at the lowest and highest ages (figure 3c). The model also suggest that excess mortality was likely higher for men compared to women across the entire range of ages (figure 3d).

**Figure 3.**
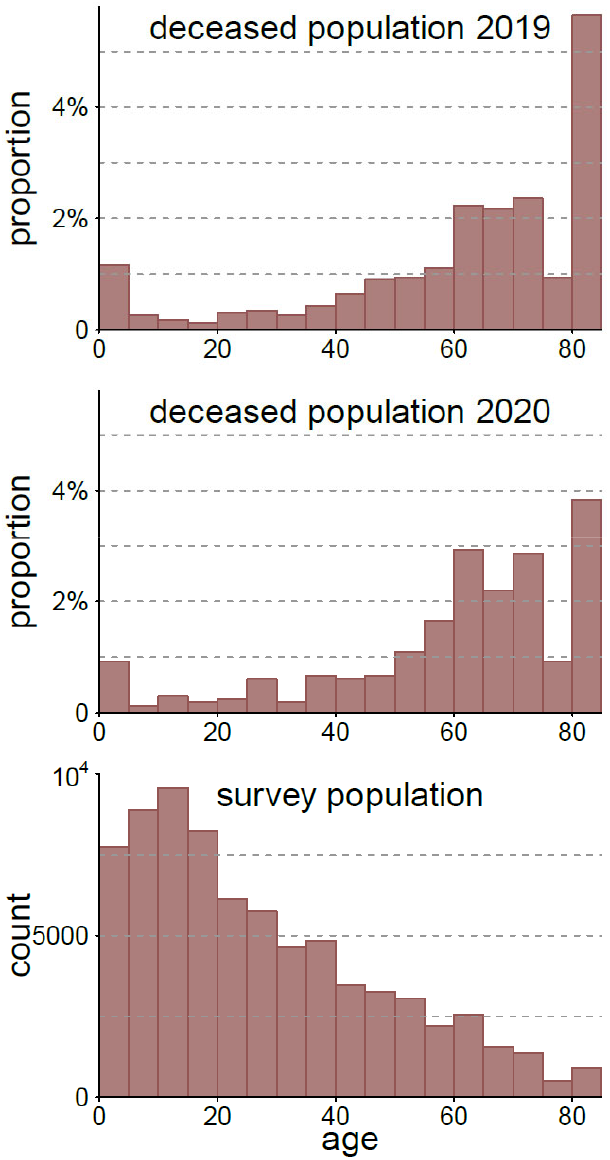
(a, b) Distribution of age at death in study households in 2019 and 2020. (c) Histogram of ages of all survey individuals in Jan 2020. Reported ages above 80 are combined.

After poststratification across gender and education, the model shows no indication of an increase in mortality rate during Feb–Oct for 2020 compared to 2019 in all age group (figure 4a). Comparing Feb–Oct 2020 with the baseline using estimate [eq_excess_m2] in Appendix A, the monthly mortality changes by -0.04 (95% CI, -0.11 to 0.04), deaths per thousand people per month averaged over all ages, and -0.12 (95% C.I., -0.52 to 0.29) at ages 50 and older. The baseline monthly baseline mortality is estimated to be 0.51 (.41–.63) for all age average, or 2.4 (1.9–3.0)) deaths per thousand per month. Hence, using estimate [eq_excess_m3] in Appendix A, our inferred mortality declines amount to -8% (−21% to +7%) or -5% (−21% to +12%) in percentage changes. This overall decline was largely due to the mortality decline in the 80+ group. Excess mortality does not vary much with household education level (a proxy for socioeconomic level) and gender.

**Figure 4.**
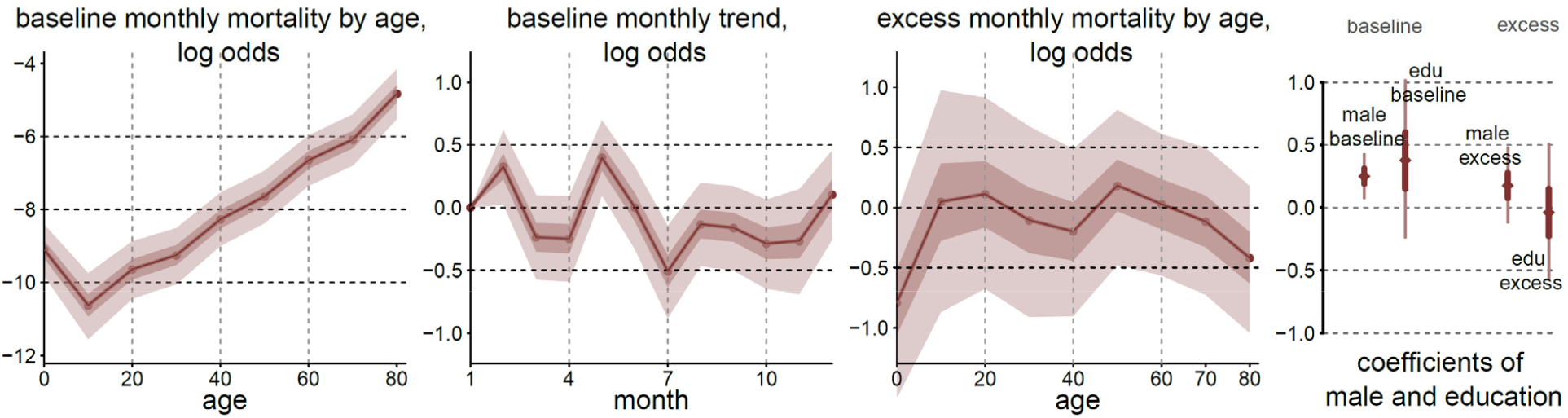
Decomposition of all modeled factors to baseline and excess mortality in log odds, visualized by the posterior mean, 50\% and 95\% confidence intervals: (a) the age-specific baseline monthly mortality log odds across ages; (b) the baseline month-of-year effect of the monthly mortality log odds across months; (c) the age-specific excess monthly mortality during February-October 2020; (d) the baseline and excess log odds associated with male or family education level.

**Figure 5.**
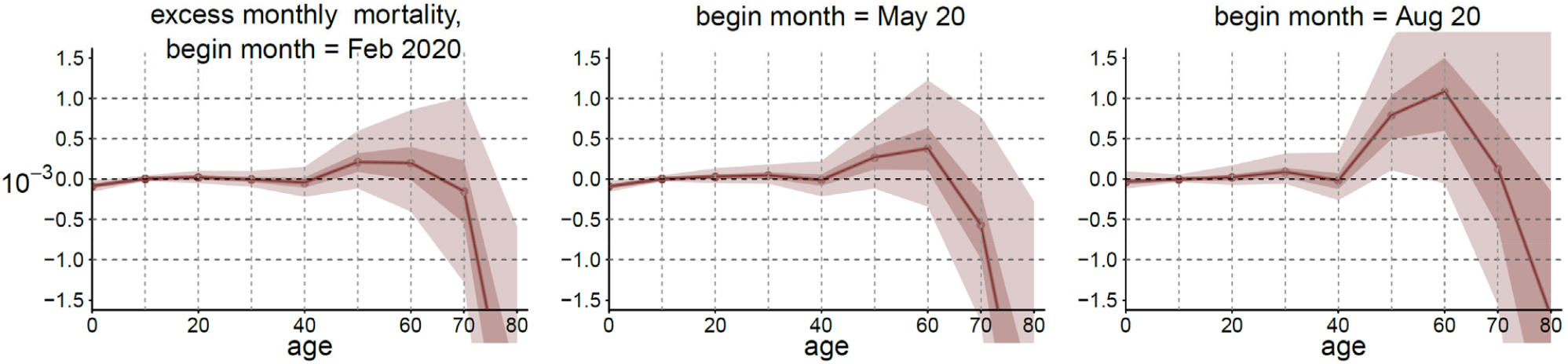
The age-specific average excess monthly mortality rate and its confidence interval. The comparison window always ends in Oct 2020 while the starting month varies from Februry 2020 to August 2020.

The excess mortality changes as the boundary between the two comparison window is shifted to later in 2020. Comparing Aug–Oct 2020 to the baseline, the model indicates that monthly mortality rate increased by 0.10 (95% CI, -0.06 to 0.30), death per thousand people per month). For age 50+, the monthly mortality increased by 0.58 (95% CI, -0.23 to 1.60). The inferred posterior mean corresponds to a 20% (25%) increase in percentage in all (50+) population, with considerable uncertainty.

The circumstances and causes of a total of 795 deaths reported for 2019–20 during the two rounds of phone calls did not vary much over time. In 2019 and 2020, respectively, 73 and 67% of deaths were preceded by consultation with a doctor or nurse, 17 and 18% of deaths occurred at a hospital, and 6.9 and 7.6% were the result of injury. Heart disease and stroke combined reportedly caused 41 and 47% of deaths in 2019 and 2020, respectively, and cancer reportedly caused 9.0 and 9.1% of deaths. The proportion of deaths attributed to lung disease actually went down from 6.4 to 3.7%. Among the 30 deaths attributed to lung disease in 2019, COVID-19 related symptoms such as fever, headache, cough, sore throat, breathing difficulty, loss of sense of smell, muscle aches, and chills were reported 64 times. Among the 12 deaths attributed to lung disease in 2020, the same symptoms were reported 30 times.

### Economic impacts

The lockdown had a large economic impact on our study population. Survey responses show that household income where the main bread earner had a salaried job declined by 40% on average in May 2020, although the decline was reduced to 30% by November 2020 (figure 6b). For self-employed bread earners in business, the decline in profit was 60% and barely recovered later in the year. Over the same period, 25% of households reported that they couldn’t obtain an essential food item in May 2020 because of reduced income, although this proportion had declined to 9% by November 2020. Even when the mobility of people reverted to the pre-pandemic levels, the negative economic impact of the pandemic was not proportionally reversed.

**Figure 6.**
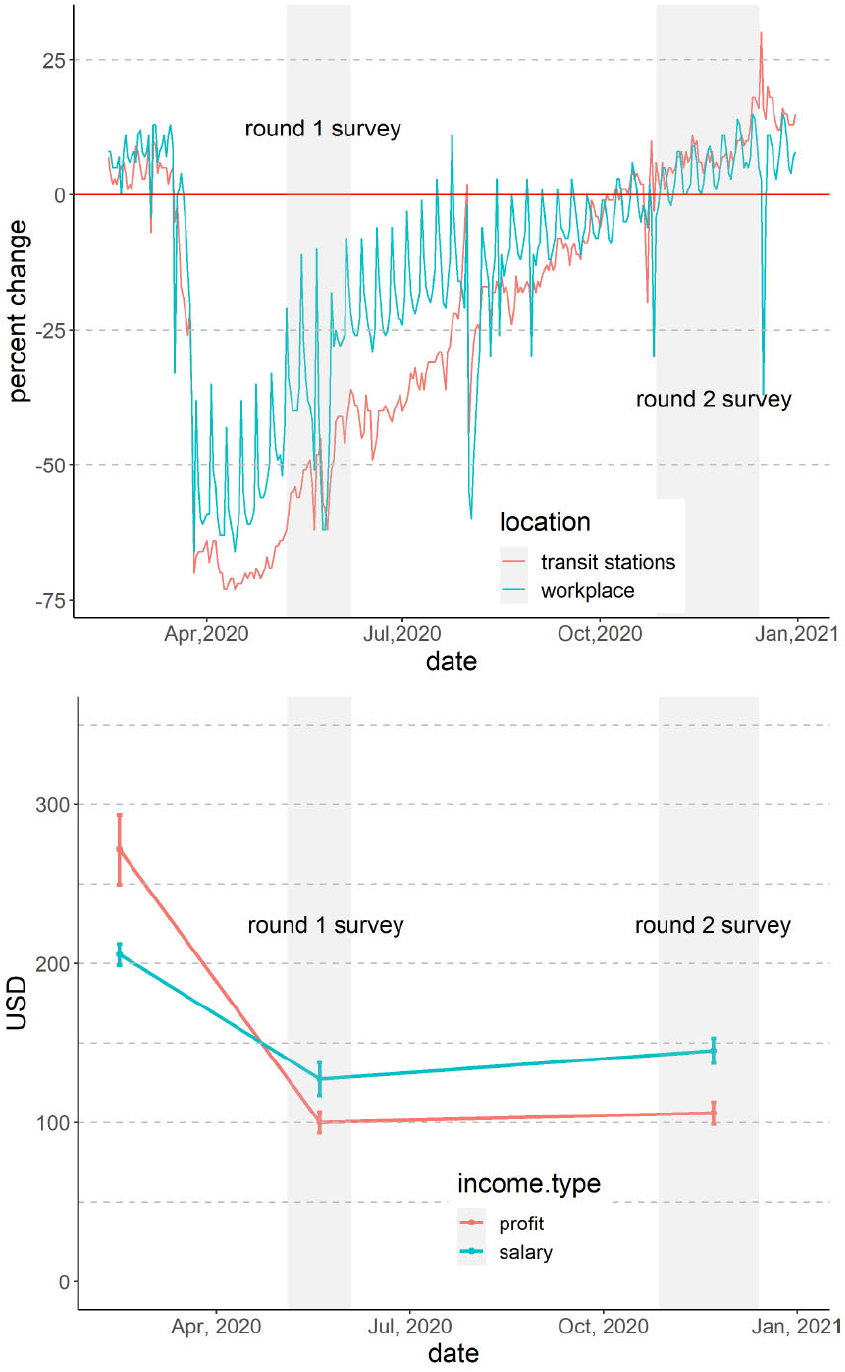
(a) Mobility change: the percentage change in mobility provided by Google COVID-19 Community Mobility Reports. The baseline mobility is the median of mobility index from January 3-February 6, 2020. (b) Income change: self-reported job salary or business profit from pre-COVID to November 2020.

## DISCUSSION

The annualized mortality rate of 6.1 per 1000 calculated over the entire 22 months covered by the study is somewhat higher than the official 2019 estimate for rural Bangladesh of 5.4 per 1000 (9). The difference is plausible given significant geographic differences in rural mortality across the country. The posterior mean of the annualized baseline mortality rates inferred from the model range from a minimum of 0.5 per 1000 for the 10–19 age range to a maximum of 150 per 1000 for 80 years and older (figure 3a). These values are also consistent with existing national statistics collected as recently as 2019 (9).

On the basis of the census data collected on three occasions, once in person and twice over the phone, we conclude that mortality did not increase in 2020 across our 135 study villages or paras. In fact, our best estimate is that mortality declined by 8%. This decrease in mortality can be potentially explained by a decline in mortality from other causes such as road accidents or the seasonal flu caused by reduced travel and social interactions, respectively. On average, the net impact of COVID-19 therefore does not appear to have come close to the levels of excess mortality of 20% and over in 2020 reported for over two dozen countries including the US (2). The 95% confidence interval obtained from the model indicates a one in forty chance that mortality actually increased by more than 7% in our study villages. Applied to the months of February through October 2020 along with an annual mortality of 820,000 for the country overall in 2019 (9), our upper bound corresponds to a one in forty chance of 43,000 additional deaths or more. The official death count attributed to COVID-19 had reached 5,900 by October 31, 2020. Combining these two figures indicates that there is only a one in forty chance that COVID-19 deaths were under-reported by more than a factor of 7. This comparison assume that both COVID-19 mortality and the reporting of COVID-19 mortality were similar in urban and rural areas, which probably was not the case. Our data could suggest that all the reported COVID-19 mortality was limited to urban areas, which account for about 1/3 of the Bangladesh population. Even under this scenario, reported COVID-19 mortality would correspond to only about of 3% or urban mortality in previous years.

We do not claim that our sample of 135 villages is representative of all of rural Bangladesh, although Table 1 provide some reassurance. Another limitation of the study is that a quarter of the households surveyed in January 2020 could not be reached over the phone by November 2020. We cannot exclude that households with a member who recently died is less likely to pick up the phone or less willing to participate in the survey. Our approach to calculate aggregate mortality using demographic group-level mortality corrects for it, to the extent that mortality estimated for a particular age, sex and education group is not biased by attrition. The repeated census approach may also not have entirely eliminated a tendency not to report a recent death, especially if it was associated with COVID-19 symptoms because of widely reported stigma, especially at the beginning of the pandemic (21). We have separate evidence from our survey that COVID-19-like symptoms were under-reported by affected households on the basis of responses from neighboring households integrated at the village and para level.

Various hypotheses have been proposed to explain the apparently lower impact of COVID-19 in some low-income countries (22). The effect of a relatively young population cannot be a factor in our study given that we are comparing the same population over two years. Spending more time outside or in well-ventilated houses has been invoked as an explanation, but another possibility is that previous infections in regions like rural Bangladesh could have dampened the symptoms of COVID-19 (23).

Mobility data and our economic impact data both indicate that the reach of the government’s interventions extended to rural areas of Bangladesh. The absence of excess mortality in our study population suggests that limiting gatherings, encouraging masks, and maintaining social distance were broadly successful in these areas in 2020 (24). At the same time, our data show that measures restricting work, trade, and travel imposed an economic burden on rural households that extended over at least six months. Similar impacts have previously been reported for 2020, including from a neighboring area of Bangladesh where both a large drop in household income and increased food insecurity were documented (25). There is therefore a critical need for the government of Bangladesh, and low-income countries more generally, to expand their social safety net to the population affected by strong non-pharmaceutical interventions to keep the COVID-19 pandemic under control (7,26).

A riskier interpretation of our results is that the de facto national lockdown imposed in March 2020 was excessive given the high economic cost and that no excess mortality was observed in our study villages. We do not endorse this view but the government of Bangladesh and other low-income countries might want to consider in the future more regionally targeted lockdowns that distinguish urban and rural areas among other factors. Such a targeted approach crucially depends, however, on monitoring across the country and, therefore, on the widespread availability of COVID-19 testing. The rapid growth in the pace testing at the beginning of the pandemic was arrested after the imposition of a charge for testing of BDT200 (US$2.40) at government facilities, a charge halved since, and BDT500 (US$6.00) for samples collected from home (27). This suggest free COVID-19 testing should be made available again throughout the country and that increased testing capacity will be needed to handle the likely surge in demand. By analogy to the impact free well-testing for arsenic had in rural Bangladesh over a decade ago (28,29), widely available COVID-19 testing could have an additional impact by encouraging health-protective behavior such as avoiding gatherings, wearing a mask, and maintaining social distance. The lessons from the pandemic in 2020 remain relevant in 2021 and may remain so for a while to come. When the number of COVID-19 cases and deaths started to increase rapidly in March 2021, the government of Bangladesh faced a difficult decision between the economic cost of a new national lockdown and the public health cost of not imposing restrictions (figure 2). A lockdown was re-imposed in early April 2021 and, unlike in neighboring India (30), the worrisome trends fortunately reversed by the beginning of May 2021.

## CONCLUSIONS

Where national statistics make this possible, excess mortality data have provided a powerful measure of the impact of COVID-19. In spite of their limitations, our repeated census surveys of a sizeable rural population fill this gap for a country where such statistics are not available. For reasons that deserve further study, it does not appear that COVID-19 had an impact on mortality in rural Bangladesh in 2020 anywhere close to that of the on-going pandemic in many other countries. While this suggests restrictions imposed by the government had the desired effect, our study also highlights the need for corrective public programs to address the very high economic burden resulting from such measures.

## Data Availability

Data and code are available at https://github.com/yao-yl/mortalityPaper. All confidential information was removed from the posted survey data.

## Acknowledgments

We thank the large teams of enumerators involved in the field and phone surveys. We particularly thank the residents of our study villages whose patience in a stressful situation was tested by a number of long phone calls. A number of colleagues including Joel Cohen (Rockefeller University and Columbia University), Mushfiq Mobarak (Yale University), Stephen Luby (Stanford University), Jeffrey Shaman (Columbia University), and Moyeen Uddin (Civil Registration and Vital Statistics Initiative, Government of Bangladesh) provided helpful advice over the course of the study.

## Contributors

PB and AvG conceived the original arsenic mitigation study and adapted it with YW, NAJ, and SR to reconstruct mortality. NAJ, SR, MAH recruited, trained, and supervised the enumerators. The data were checked for errors and compiled by NAJ, SR, and YW. YY conducted the statistical analysis with input from PB, YW, NAJ, and AvG. AvG, YY, and PB drafted the manuscript, which was edited based on input from all co-authors.

## Ethics approval

Institutional Review Boards at both Michigan State University and Columbia University approved the study.

## Appendix A: Model details

### Baseline mortality rate

We use the following notation for the four discrete variables: month *j*_1_ = 1, …, 22; age during the evaluation month, *j*_2_ = 1, …, 9; gender *j*_3_ = 1, 2; and household education *j*_4_ = 1, 2. Any individual stratum can be written as *j* = (*j*_1_, *j*_2_, *j*_3_, *j*_4_). In each stratum *j*, we count *n*_*j*_ the number of surveyed individuals that were alive in the beginning of the month and accessible throughout the month, and *y* _*j*_ the number of deceased individuals. Assuming independent sampling, the data model is

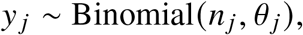

where the parameter *θ*_*j*_ is what we want to estimate: the monthly mortality rate in month *j*_1_ for age group *j*_2_, gender *j*_3_, and household education level *j*_4_.

The baseline (*j*_1_ *< t*_start_) mortality rate for stratum *j* is modeled as a function of month and individual’s age-sex-education attributes,

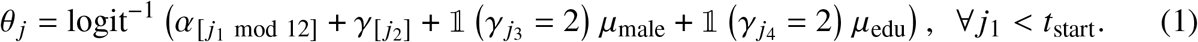

where the free parameters are

- The month-of-year factor, or the seasonal trend, denoted by *α*_1_, …, *α*_12_ for January to December each year. Due to periodicity, cell *j* processes this seasonal factor 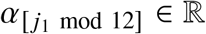. For identification, we set January as baseline such that *α*_1_ = 0.
- The age factor, denoted by *γ*_1_, …, *γ*_9_ for 9 age categories. The age factor for cell *j* is 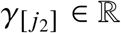.
- The male factor *µ*_male_ ∈ ℝ. We set female as reference, then the sex factor for cell *j* is 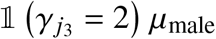.
- The education factor *µ*_edu_ ∈ ℝ. We set non-education as reference, and the education factor for cell *j* is 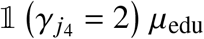.

*Mortality in 2020*. We take a flexible approach by specifying a comparison-starting month *t*_start_, which splits time into the two periods (a) the baseline: Jan 2019, Feb 2019, …, *t*_start_ − 1, and (b) the potential excess period: *t*_start_, *t*_start_ + 1, …, Oct 2020. During the second period, on top of the baseline model (1), we model the stratum-specific excess risk:

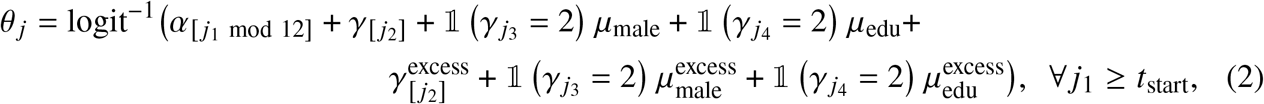

where additional parameters *γ*^excess^, 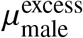 and 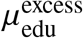 represent the excess risk in 2020 associated in the age, sex, and education, on top of the baseline risk.

### Partial-pooling and priors

We complete the model by weakly-informative priors:

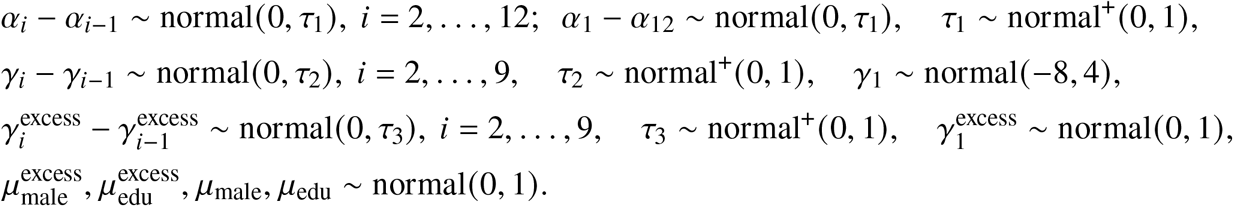

### Aggregate mortality in the population

After model is fitted using Markov chain Monte Carlo simulations, we generate posterior simulations of the demographic-specific baseline mortality :

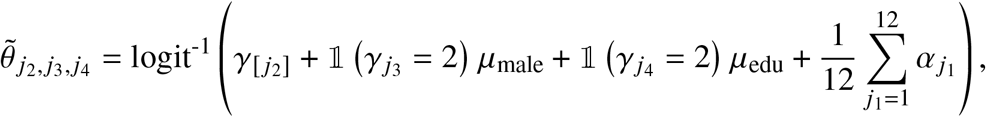

and the demographic-specifc mortality in 2020 after *t*_start_:

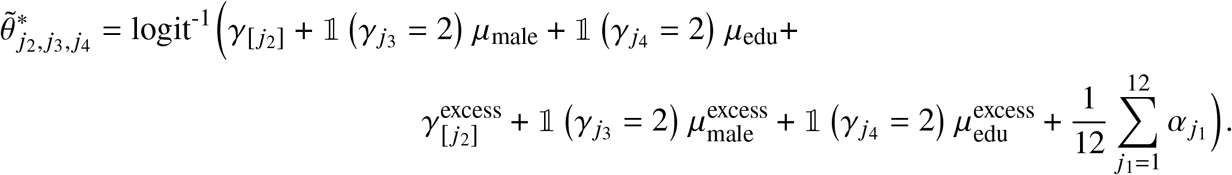

We use census data to construct the demographic post-stratification weight 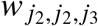 : the propor-tion of individuals in our round-1 survey with age *j*_2_, gender *j*_2_ and educational level *j*_4_. Then the monthly excess mortality of any given age group *j*_2_ is computed by

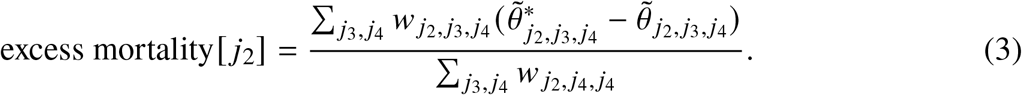

Likewise, we aggregate this excess mortality across all ages in the population:

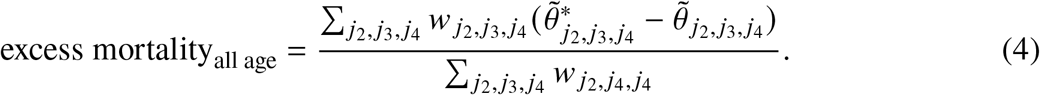

To compute the relative mortality change, we divide this excess rate by the average baseline mortality,

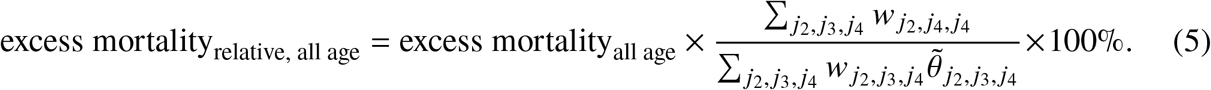

## Appendix B: Additional graphs

**Figure 7:**
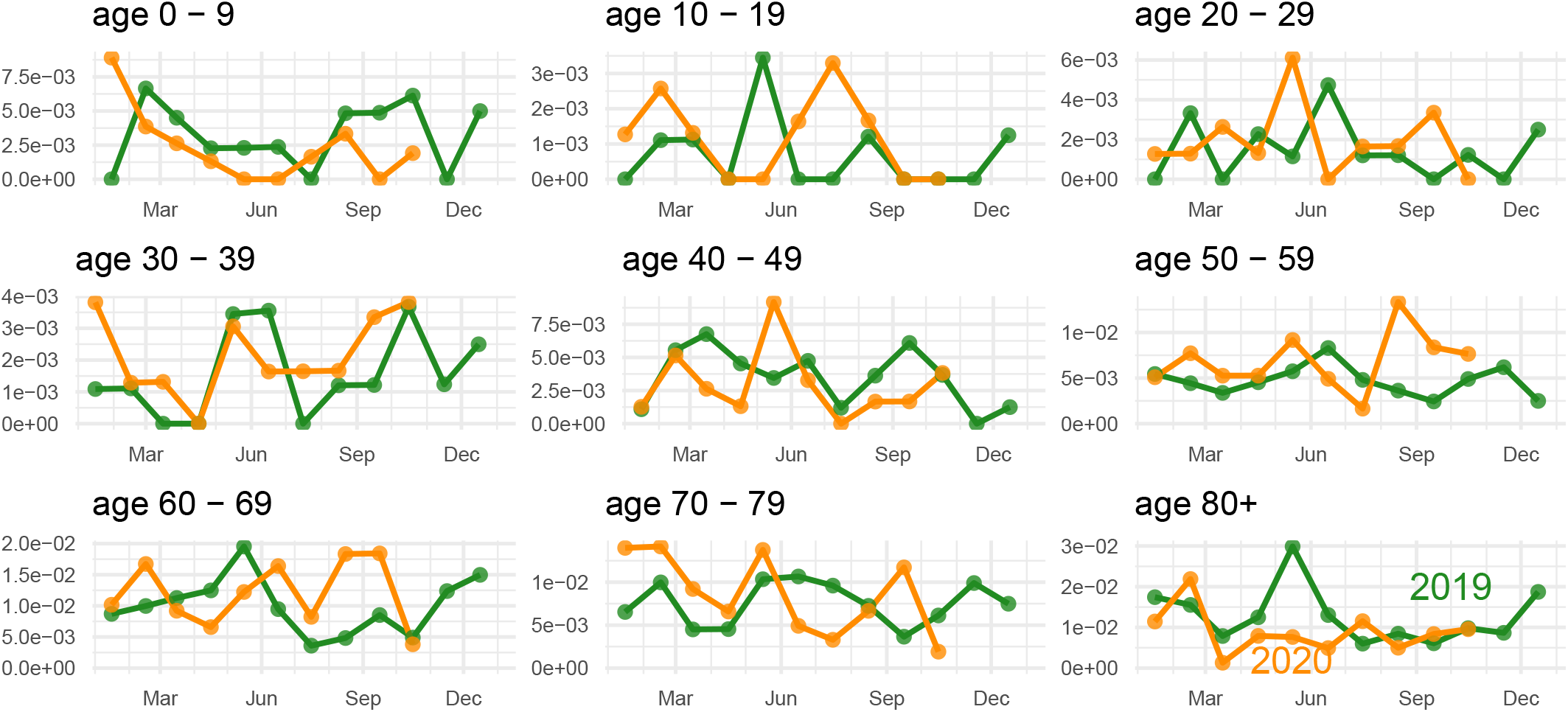
The empirical monthly mortality rate by age in 2019 (green line) and 2020 (orange). The mortality decline during Feb–Oct 2020 was evident for age group 0–9 and 80+.

**Figure 8:**
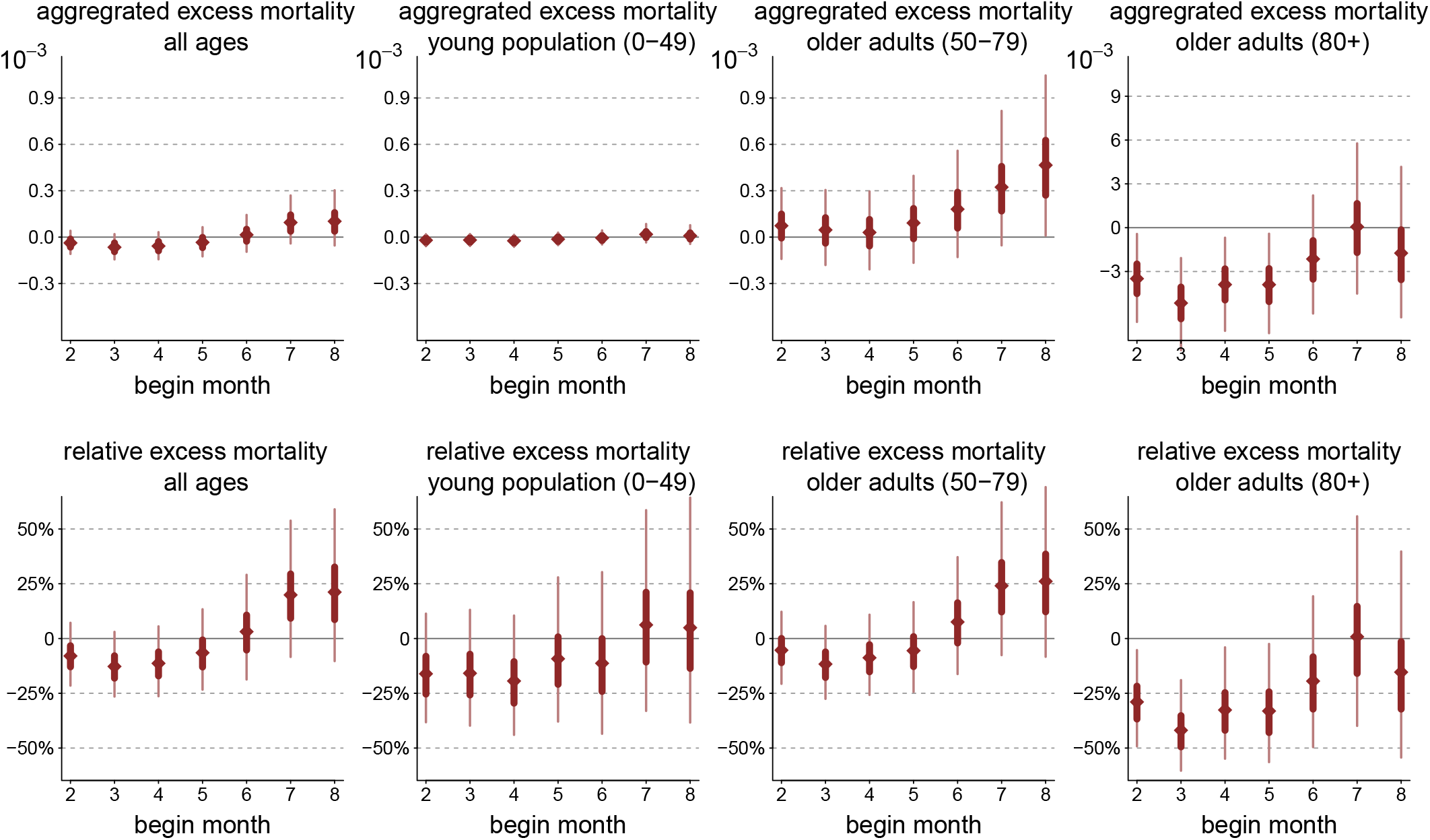
The posterior inference of the age aggregated (all ages, 0–49, 50–79, and 80+) monthly excess mortality rates and confidence intervals. The upper row is absolute change compared with baseline (extra death per thousand per month), and the lower row is the relative changes in percentage. The starting month of the comparison window *t*_start_ varies from Feb 2020 to Aug 2020.

**Figure 9:**
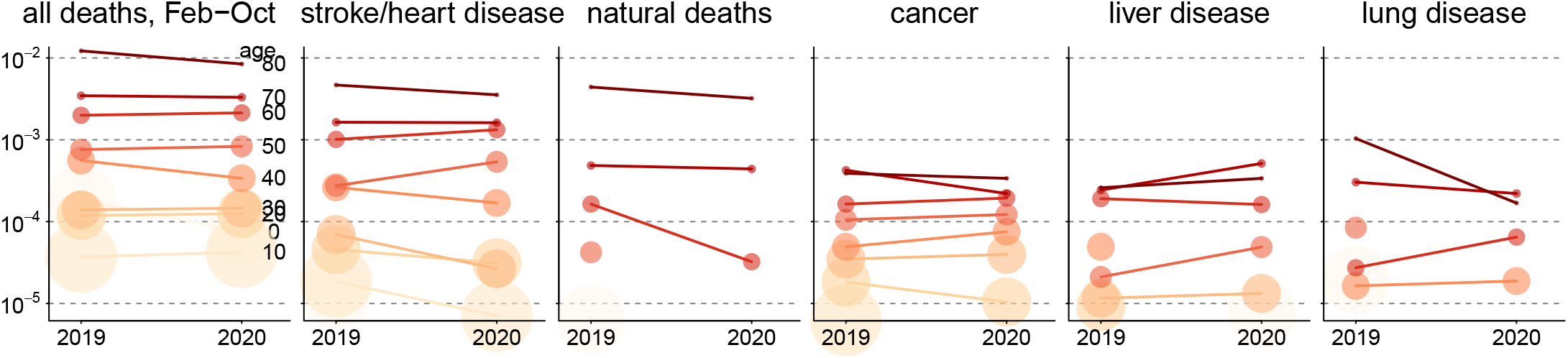
The empirical comparison of the age-specific proportion of survey samples (among all survey objects) who (1) died from all causes, (2) died from “stroke/heart discese”, (3) died from from “natural death”, (4) died from “cancer”, (5) died from “liver disease”, (6) died from “lung disease”. The comparison is between Feb–Oct 2019 and Feb–Oct 2020. The dot sizes are the accessible population of that age group in the survey.

**Figure 10:**
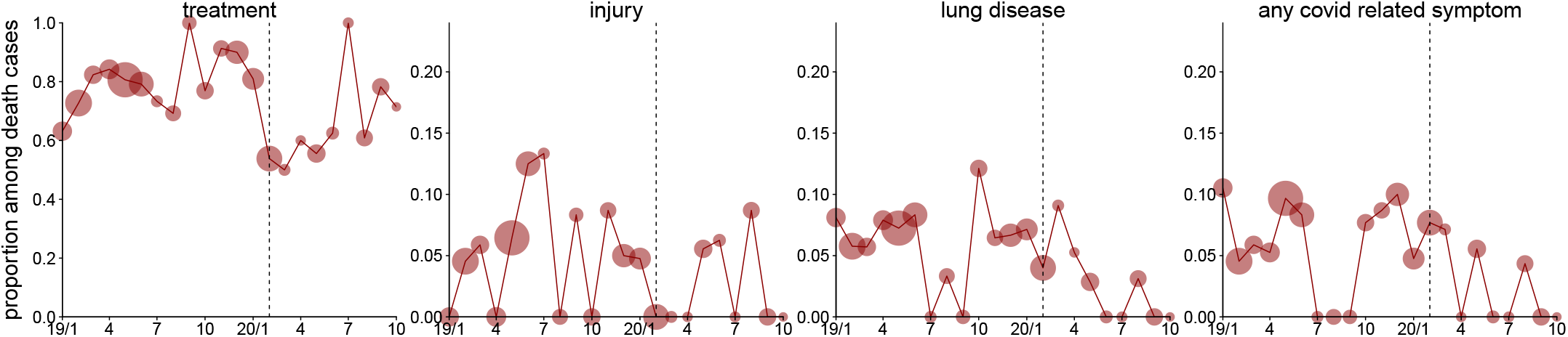
The empirical percentage of surveyed death case (among all deaths) of who (a) received any medical treatment, (b) were injured, (c) went through lung disease, and (d) went through any one of the COVID-related symptoms (headache, muscle aches, chill, cough, sore throat, lose-of-sense-of-smell, breathing difficulty, and fever). The dot size indicates the total death cases during each month. The category (c) and (d) largely overlap, while none of them exhibited an increasing trend in 2020.

## REFERENCES

1. Beaney T, Clarke JM, Jain V, Golestaneh AK, Lyons G, Salman D, et al. Excess mortality: The gold standard in measuring the impact of COVID-19 worldwide? Journal of the Royal Society of Medicine. 2020;113(9):329–34.

2. Karlinsky A, Kobak D. The world mortality dataset: Tracking excess mortality across countries during the COVID-19 pandemic. medRxiv. 2021; doi: 10.1101/2021.01.27.21250604

3. Setel PW, Macfarlane SB, Szreter S, Mikkelsen L, Jha P, Stout S, et al. A scandal of invisibility: Making everyone count by counting everyone. Lancet. 2007;370(9598):1569–77.

4. Mikkelsen L, Phillips DE, AbouZahr C, Setel PW, De Savigny D, Lozano R, et al. A global assessment of civil registration and vital statistics systems: Monitoring data quality and progress. The Lancet. 2015;386(10001):1395–406.

5. Chatterjee P. Is India missing COVID-19 deaths? Lancet. 2020;396(10252):657.

6. Checchi F, Roberts L. Documenting mortality in crises: What keeps us from doing better? PLoS Med. 2008;5(7):e146.

7. Egger D, Miguel E, Warren SS, Shenoy A, Collins E, Karlan D, et al. Falling living standards during the COVID-19 crisis: Quantitative evidence from nine developing countries. Science Advances. 2021;7(6):eabe0997.

8. World Health Organization. WHO COVID-19 dashboard [Internet]. 2021. Available from: {https://covid19.who.int}

9. Bangladesh Bureau of Statistics. Report on sample vital registration system (SVRS) 2019 [Internet]. 2020. Available from: {https://drive.google.com/file/d/1TtdcJaDyc7vf5u7Aza8GmqqGv8eqP6JN/view}

10. Uddin M, Ashrafi SAA, Azad AK, Chowdhury A, Chowdhury HR, Riley ID, et al. Improving coverage of civil registration and vital statistics, Bangladesh. Bulletin of the World Health Organization. 2019;97(9):637.

11. Elyazar IR, Surendra H, Ekawati L, Djaafara BA, Nurhasim A, Hidayana I, et al. Excess mortality during the first ten months of COVID-19 epidemic at Jakarta, Indonesia. medRxiv. 2020; : https://doi.org/10.1101/2020.12.14.20248159.

12. Besson ESK, Norris A, Ghouth ASB, Freemantle T, Alhaffar M, Vazquez Y, et al. Excess mortality during the COVID-19 pandemic: A geospatial and statistical analysis in Aden governorate, Yemen. BMJ Global Health. 2021;6(3):e004564.

13. Biswas RK, Afiaz A, Huq S. Underreporting COVID-19: The curious case of the Indian subcontinent. Epidemiology & Infection. 2020;148.

14. Mukherjee S. Why does the pandemic seem to be hitting some countries harder than others? The New Yorker.; March 1, 2021.

15. Hygiene London School of, Medicine Tropical. The use of epidemiological tools in conflict-affected populations: Open-access educational resources for policy-makers. [Internet]. 2009. Available from: {http://conflict.lshtm.ac.uk/page_02.htm}

16. Bangladesh Bureau of Statistics. Household income and expenditure survey (HIES). 2016.

17. Gelman A, Little TC. Poststratification into many categories using hierarchical logistic regression. Survey Methodology. 1997;23:127–35.

18. Park DK, Gelman A, Bafumi J. Bayesian multilevel estimation with poststratification: State-level estimates from national polls. Political Analysis. 2004;375–85.

19. Stan Development Team. Stan user’s guide [Internet]. 2021. Available from: {https://mc-stan.org}

20. Becker S, Weng S. Seasonal patterns of deaths in Matlab, bangladesh. International journal of epidemiology. 1998;27(5):814–23.

21. Mahmud A, Islam MR. Social stigma as a barrier to COVID-19 responses to community well-being in Bangladesh. International Journal of Community Well-Being. 2020;1–7.

22. De Weerdt J, Van Damme W. Health, wealth and the double paradox of COVID-19 mortality in low-income countries. 2021. Available from: {https://ssrn.com/abstract=3793427}

23. Cepon-Robins TJ, Gildner TE. Old friends meet a new foe: A potential role for immune-priming parasites in mitigating COVID-19 morbidity and mortality. Evolution, Medicine, and Public Health. 2020;2020(1):234–48.

24. Ge Y, Zhang W, Liu H, Ruktanonchai CW, Hu M, Wu X, et al. Effects of worldwide interventions and vaccination on COVID-19 between waves and countries. medRxiv 2021033121254702. 2021;

25. Hamadani JD, Hasan MI, Baldi AJ, Hossain SJ, Shiraji S, Bhuiyan MSA, et al. Immediate impact of stay-at-home orders to control COVID-19 transmission on socioeconomic conditions, food insecurity, mental health, and intimate partner violence in Bangladeshi women and their families: An interrupted time series. The Lancet Global Health. 2020;8(11):e1380–9.

26. Gerard F, Imbert C, Orkin K. Social protection response to the COVID-19 crisis: Options for developing countries. Oxford Review of Economic Policy. 2020;36(Supplement_1):S281–96.

27. Cousins S. Bangladesh’s COVID-19 testing criticised. The Lancet. 2020;396(10251):591.

28. Madajewicz M, Pfaff A, Geen A van, Graziano J, Hussein I, Momotaj H, et al. Can information alone both improve awareness and change behavior? Arsenic contamination of groundwater in Bangladesh. Journal of Development Economics 2007, vol. 84, issue 2, 731–754.

29. Huhmann BL, Harvey CF, Navas-Acien A, Graziano J, Parvez F, Chen Y, et al. Changes in arsenic exposure in Araihazar, Bangladesh from 2001 through 2015 following a blanket well testing and education campaign. Environment International. 2019;125:82–9.

30. Mallapaty S. India’s massive COVID surge puzzles scientists. Nature. 2021; https://www.nature.com/articles/d41586-021-01059-y

